# Measuring the exposure of Black, Asian and other ethnic groups to Covid-infected neighbourhoods in English towns and cities

**DOI:** 10.1101/2021.03.04.21252893

**Authors:** Richard Harris, Chris Brunsdon

## Abstract

Drawing on the work of The Doreen Lawrence Review – a report on the disproportionate impact of Covid-19 on Black, Asian and minority ethnic communities in the UK – this paper develops an index of exposure, measuring which ethnic groups have been most exposed to Covid-19 infected residential neighbourhoods during the first and second waves of the pandemic in England. The index is based on a Bayesian Poisson model with a random intercept in the linear predictor, allowing for extra-Poisson variation at neighbourhood and town/city scales. This permits within-city differences to be decoupled from broader regional trends in the disease. The research finds that members of ethnic minority groups tend to be living in areas with higher infection rates but also that the risk of exposure is distributed unevenly across these groups. Initially, in the first wave, the disease disproportionately affected Black residents. As the pandemic has progressed, especially the Pakistani but also the Bangladeshi and Indian groups have had the highest exposure. This higher exposure of the Pakistani group is not straightforwardly a function of neighbourhood deprivation because it is present across a range of average house prices. However, we find evidence to support the view, expressed in The Doreen Lawrence Review, that it is linked to occupational and environmental exposure, particularly residential density.

## 1. Introduction

> Covid-19 is having a disproportionate and devastating impact on ethnic minority communities. Not only are Black, Asian and minority ethnic people dying at a disproportionate rate, they are also overexposed to the virus and more likely to suffer the economic consequences (*The Doreen Lawrence Review*, 2020: opening sentences of the Executive Summary, emphasis added).

This paper looks at the exposure of Black, Asian and minority ethnic (BAME) groups to neighbourhoods of greater than expected rates of Covid-19 infection, in English major towns and cities, over the first 11 months of the pandemic. It takes, as its stating point, the findings of *The Doreen Lawrence Review* (2020), which explores the disproportionate impact of the current pandemic on BAME groups. The report’s claim that “this virus has exposed the devastating impact of structural racism” in Britain appears to be supported, as a matter of life and death, by mortality statistics from the Office for National Statistics (ONS, 2020c). These show that the rate of death involving coronavirus among Black African males was 3.8 times higher than for those of White background; for Black African females it was 2.9 times higher. All groups other than the Chinese had a statistically significant higher rate of mortality than those of a White ethnicity.

However, the ONS data are for deaths only up to July 28, 2020. Similarly, *The Doreen Lawrence Review*, is informed by data from the first half of that year. The timing of the data matters because there have been regional and sub-regional geographies to the disease that have changed as the disease has spread, contracted and spread again across the UK. The shifting geography is why, at various stages of the pandemic, some places have faced greater restrictions than have others on people mixing and on businesses trading – the ‘local lockdowns’ and tiered system of place-based restrictions.

The spatio-temporal variations in the disease would be of less relevance if the prevalence of various BAME groups was random or uniform across the country. It is not. Consequently, the demographic characteristics of the at-risk population are changing with the geography of the disease. At the beginning of the pandemic, it was disproportionately found in London, a geographical attribute that goes some way to explaining the higher impact upon Black communities in those early months (Harris, 2020). Later on, it was more greatly affecting Asian groups in places such as Leicester.

This paper looks at neighbourhood levels of infection over the period from the seven days ending March 21, 2020, to the seven ending February 19, 2021. It is a period over which the initial wave of the pandemic rose, spread and then contracted into more regionalised clusters of infection during the spring and summer of 2020, spreading nationally again in a second, autumn and winter wave. The second led to more infections than the first, driven by a more infectious, mutated strain of the virus that emerged towards the end of the year and into 2021. The hypothesis is that BAME groups will have continued to be overexposed to the disease because the occupational and environmental exposures raised in *The Review* will have persisted. However, this is conjecture until it is evidenced.

To provide the evidence, a Bayesian Poisson model is developed to identify, for each week of the data, the neighbourhoods in English major towns and cities that have the highest rates of infection. The model allows neighbourhoods to be identified that have the highest prevalence of the disease relative to the national and to their immediate context. The question is then one of who lives in those neighbourhoods: is it disproportionately members of one or more BAME groups? If so, do the inequalities in exposure persist over time? To answer this, an index of exposure is developed, based on the type found in studies of ethnic segregation but here measuring the relative exposure of each ethnic group to neighbourhoods of Covid infections.

The research finds that BAME groups do tend to be living in areas with higher infection rates but their exposure to these neighbourhoods is unevenly distributed across the groups. It is especially the Pakistani but also the Bangladeshi and Indian groups that have had the highest exposures for most weeks. Those higher values, for the Pakistani group, remain evident when the index is stratified into deciles based on local property prices, suggesting they are not simply a function of neighbourhood deprivation. Environmental exposure, measured as residential density (people per dwelling), appears to be the main exploratory factor, with occupational exposure also a contributory factor.

## 2. Literature Review and Context

The *Doreen Lawrence Review*, cited in the Introduction, was published by the UK Labour Party as a response to the Covid-19 pandemic in England, looking at structural inequalities in who is infected and dying from the disease. It argues that “Black, Asian and minority ethnic people have been overexposed, under protected, stigmatised and overlooked during this pandemic” (Foreword to the Review). At about the same time, a report by the Institute for Public Policy Research (IPPR) and Runnymede Trust described the extra risk of death in minority ethnic groups as “one of the starkest health inequalities in recent times” (Patel et al., 2020). It estimated that “over 58,000 and 35,000 additional deaths from Covid-19 would have occurred if the white population had experienced the same risk of death from Covid-19 as the black and south Asian populations respectively [over the period between March and May 2020].” Those numbers would near-double or more the actual 38,122 deaths. At the end of November 2020, Channel 4, one of the five main terrestrial channels in the UK, broadcast a documentary, entitled ‘*Is Covid Racist?*’, picking-up on occupational exposure in the medical profession and asking, “why so many Black, Asian and Minority Ethnic NHS colleagues have died from Covid-19” (https://www.channel4.com/programmes/is-covid-racist).

The impression that BAME groups are at greater risk is supported by the data. Although published by a political party, those found in *The Review* are not partisan but are taken from and/or support the findings of other studies, primarily from the first half of 2020. Early analysis, published by Public Health England in June 2020, showed that Black ethnic groups were, at that time, most likely to be diagnosed with Covid-19 and that death rates were highest among Black and Asian groups. It observed that “this is the opposite of what is seen in previous years, when the mortality rates were lower in Asian and Black ethnic groups than White ethnic groups” (Public Heath England, 2020: 6).^1^

The analysis by Public Health England covers the first wave of the disease in England. The first deaths were at the end of February / early March 2020, with the daily deaths in that wave peaking on April 21, with 1,172 deaths. Sadly, that number was exceeded for much of January 2021.^2^ At first, London had the highest death rate of any English region. This is linked to the higher rates for, especially, Black but also Asian groups, because these groups are disproportionately resident in the capital: according to the most recent Census, 59 per cent of the entire Black residential population of England lived in London (on March 27, 2011) and 36 per cent of the Asian population. In comparison, 15 per cent of the English population were London residents. However, it is not simply a ‘London effect’. If it was, it would apply equally to all ethnic groups who live there, whereas, within London, areas with higher percentages of BAME groups also had higher death rates (Harris, 2020).

As, through the summer and early autumn, attention moved to towns and cities in the Midlands, North East and North West of England – regions within which local lockdown regimes were put in place to curb rising infections after the first national lockdown had ended – ethnic inequalities still persisted but had shifted more to Asian groups. When the period 20-26 August 2020 is compared with 28 May to 3 June, the number of tests had increased by 83 per cent for Asian/Asian British groups, by 44 per cent for White groups but by only 28 per cent for Black/Black British groups (Department of Health & Social Care, 2020: Table 2). This reflects the spread of Covid-19 outside of London, in places such as Oldham, Bradford, Blackburn, Manchester, Rochdale, Northampton and Leicester, where there are more of the Asian groups. Analysis by the Office for National Statistics (ONS, 2020b: Figure 8) showed that Covid-19 had a proportionally higher impact on the age-standardised mortality rates in the most deprived areas of England throughout the period March to July, 2020, many of which are occupied by BAME groups but not only BAME groups.

In England, the main predictors of Covid-19 vulnerability have been identified as the proportions of the population, (a) living in care homes, (b) admitted to hospital in the past five years for a long-term health condition, (c) from an ethnic minority background, and (d) living in overcrowded housing (Daras et al., 2020). The co-linearity of these variables with each other, as well as with age and occupation, could explain why income deprivation was not found to be statistically significantly in this study (that is, the effects of income deprivation were measured through the other variables). Occupations with increased risk of exposure include frontline medical staff, the emergency services, public transit workers, teachers and those working in the hospitality industry, many of which – for example, pharmacists, dental and medical practitioners, and bus drivers – have a disproportionate percentage of their workforce from BAME backgrounds (ONS, 2020d).

In the United States, the Centers for Disease Control and Prevention observe that “[s]ome of the many inequities in social determinants of health that put racial and ethnic minority groups at increased risk of getting sick and dying from Covid-19” include: discrimination; healthcare access and utilisation; occupation; educational, income and wealth gaps; and housing (CPD, 2020) – concerns that echo those found in *The Doreen Lawrence Review*. A systematic review of 50 studies, 42 from the United States of America and 8 from the United Kingdom, confirms that individuals from Black and Asian ethnicities had a higher risk of Covid-19 infection compared to White individuals (Sze et al., 2020). However, a report by the UK Government Equalities Office and Race Disparities Review (2021) stresses that “that ethnic minorities should not be considered a single group that faces similar risk factors in relation to Covid-19. Different ethnic groups have experienced different outcomes during both waves of the virus.” The analysis that is presented in this paper also supports that view.

## 3. Analytical Approach

### 3.1 About the infections data

The main dataset that is used in this paper is from the official UK Government data dashboard, https://coronavirus.data.gov.uk. Here we focus on cases of Covid-19 infection rather than deaths because only the infection data are available at weekly intervals and at a neighbourhood scale in England, allowing geographical and temporal patterns in the data to be considered. However, a data set giving the number of deaths due to Covid for each of the months from March 2020 to January 2021 was published at the same neighbourhood scale at the end of February 2021, as this research was being completed, having not previously been updated since August 2020 (ONS, 2021). We use these data to underscore that what is seen in the infection data is evident in the morality data too.

The neighbourhood geography is the Middle Level Super Output Areas (MSOAs). These are the third tier of the Census geography for England and Wales; third when aggregating upwards from the smallest, which are Output Areas. They represent a formal, administrative specification of neighbourhood but they are not arbitrary. They were designed with the criteria of broadly equal population size, socio-economic homogeneity based on accommodation type and tenure, and spatial compactness of the zones (Cockings et al., 2011). Matching them to the Office for National Statistics mid-2019 population estimates suggests an average population size of 8,605 in English major towns and cities, with an interquartile range from 7,154 to 9,640 persons.^3^

The infections data tally, by date reported, the number of individuals who had at least one positive Covid-19 test result, over that and the preceding six days. The data used in this paper span the week to March 21, 2020 to the week to February 19, 2021. The data include only pillar 1 cases until 2 July, from when pillar 2 cases also are included. Pillar 1 cases are “diagnostic tests done in Public Health England labs and hospitals for health and care workers and patients who are seriously ill” (Full Fact, 2020). Pillar 2 cases “are also diagnostic tests but done by commercial partners for the wider population. These tests are done at regional test sites, mobile testing units, satellite test centres and via home tests.”

Not everyone who has the disease is tested or necessarily displays symptoms so the numbers are underestimates of the true prevalence of the disease within the population. Especially this will be true prior to the beginning of July 2020 when it is, in effect, a count of the number hospitalised by Covid-19. The undercount is compounded throughout the study period by the suppression of the exact number in any MSOAs that had 0 to 2 cases that week. For this study, these are treated as zero values but they could be one or two.

A further problem is that test results are necessarily a function of testing – of where people are being tested and their ability to access a test. The availability of tests has changed over time. The 7-day average numbers of tests at the beginning of the months from May 2020 to February 2021 were 67,512 (May), 92,716 (June), 112,001 (July), 154,151 (August), 187,287 (September), 255,302 (October), 284,306 (November), 311,602 (December), 425,064 (January), and 6444,440 (February) (https://coronavirus.data.gov.uk/details/testing/). Although the increase is a response to the second wave of the disease, it also reflects increased capacity to test, which, in turn, allows for more positive diagnoses. That testing capacity has been constrained for some of the period of this study, with the British Medical Journal publishing a briefing in September entitled *What’s going wrong with testing in the UK?* (Wise, 2020). It is possible that the relative resilience of London to growing numbers of cases during the initial emergence of the second wave in England – from the end of September / early October, 2020 – was in part caused by insufficient testing. However, the nature of the regional economy and the type of jobs that are more easily adapted to home working may also be a factor (Harris & Cheshire, 2020).

Not everyone who has been in close contact with an infected person has necessarily been tested. On-going problems with the track-and-trace system (Triggle, Schraer & Kemp, 2020) meant about half of those who have been in contact with an infected person had not been contacted, so could harbour the disease with neither symptoms nor knowledge. There have also been changes to the data, first to harmonise them and to remove duplicates, and, secondly, to more accurately assign people to their current residential address. This raised the numbers around Universities as students were located to their term-time address, which may differ from the one on their NHS record because the latter can be a parental address if they are registered to a GP ‘back home’.

Nevertheless, the analysis assumes that tests follow symptoms, as well as the identification of known clusters of the disease, and that, therefore, the data track the disease’s spread amongst the population sufficiently well to be broadly representative of who has been infected and where they live. They are neither a random sample nor a complete census of all those who have been infected each week. However, they are the official source of infection data in the UK, used to inform public health policy and to guide preventative measures such as national and local lockdowns. We do not dispute the possibility of bias but note that the most likely systematic problem is an undercount of those who could be infected but either can least afford to socially isolate from out-of-home employment for the quarantine period required, or to travel to access a test. If such a bias exists, then it is likely to downplay the infection rate amongst BAME groups because of the intersections of ethnicity and social disadvantage, which means differences in exposure between BAME groups and the White British will be underestimated in the analysis.

### 3.2 Geography of the study

The study is confined to English major towns and cities, as defined in the ONS file of December 2015.^4^ This is based on the built-up areas geography developed following the 2011 Census (ONS, 2013). Rural and semi-urban areas are not considered, partly because of their greater population sparsity that, all things being equal, reduces person-to-person contact and therefore the transmission of Covid-19. Much of the South West region of England, for example, has had consistently lower rates of infection throughout the pandemic and is mainly rural.

However, the main reason is demographic, informed by the ethnic geography of the country. Whereas many English towns and cities are ethnically diverse, more rural areas are typically not, containing far fewer of the BAME than White population. According to the 2011 Census, 61 per cent of the White British population resides outside the major towns and cities. The next largest percentage is equal for the White Other and Mixed ethnicity groups, of whom 30 per cent do not reside in these towns and cities. For the Black Caribbean and Black African groups, it is only 11 per cent. Nationally, lower Covid-19 infection and death rates amongst the White British population reflect rural-urban patterns of living. To compare ‘white’ rural hamlets with multicultural urban settlements is problematic; they are very different types of places. Consequently, we prefer the direct comparison, asking whether differential rates of exposure remain evident between different ethnic groups, within urban settlements.

In total, 109 English towns and cities are included in the study. The smallest is Walsall, with a mid-2019 estimated population of 65,928. The largest is London, with a population of 8,924,265. Because London is so much larger than the other settlements – its population is over 35 times greater than the average of 250,479 and approaching eight times greater than the second largest settlement, Birmingham, with 1,153,804 – it is split into its 32 local authorities for the analysis: the 32 London Boroughs, with the City of London merged with Westminster. This gives a total of 140 urban ‘places’.

### 3.3 Method of estimating the relative exposure of ethnic groups to neighbourhoods of Covid-19 infection

For the analysis, an index of exposure is formed to measure how much members of the various ethnic groups are exposed, by residence, to neighbourhoods of Covid-19 infection for each week of the study period. This index is based on the residuals from a Bayesian Poisson model, which are used to identify neighbourhoods that have more or less than the expected number of Covid-19 cases, relative to: (a) the overall infection rate that week; or, (b), also relative to the town or city in which the neighbourhood is situated.

The underlying model is,

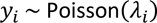

where

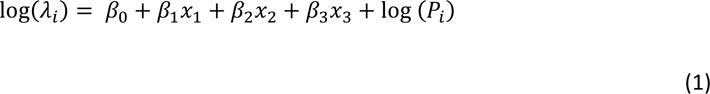

Here, *y*_*i*_ is the expected number of infection counts in MSOA, *i*, conditional on the control variables (*x*_1_, *x*_2_ and *x*_3_,). These control variables are the ratio of care home beds in the MSOA to the adult population, the percentage of the adult population aged 18 to 21, and the percentage aged 22 to 35. The first two of these variables are included to control for two distinct types of setting where infection rates have been unusually high for at least some of the pandemic but where the inhabitants of those settings do not necessarily reflect the population characteristics of the wider neighbourhood. These are care homes and student halls of residence.

The third variable, the percentage of the adult population aged 22 to 35, is included because of a period, after the first national lockdown, when infection cases were growing amongst younger adults, and because BAME groups are, on average, younger than the White British. There is a risk that this control ends-up understating the exposure of BAME groups to infected neighbourhoods if controlling for age also partially controls for ethnicity. However, we judge this preferable to potentially confusing an age effect with an ethnic one. From an analytical point of view, if observed differences remain between the ethnic groups despite potentially biasing those differences downwards then that strengthens the argument that those differences are of substantive interest.

The control variables are standardised into *z-*values in the model (units of standard deviation from their mean). No further variables are included in the model because its purpose is not to explain the variations in the infection rates but to identify them, at the neighbourhood and town/city/London Borough scales. It is adopted to enable a multilevel approach that permits the differences *between* the urban places to be separated from the differences *within* them. It is, in effect, a null model, albeit one that controls for a few distinct circumstances that are unusual and would otherwise ‘inflate’ the number of cases in their MSOA.

Returning to Equation 1, *P*_*i*_ is the number of the adult population in the MSOA and log (*P*_*i*_) is an offset (it has a prescribed coefficient of one) that gives the log of the conditional rate of infection. This is seen by re-arranging the equation,

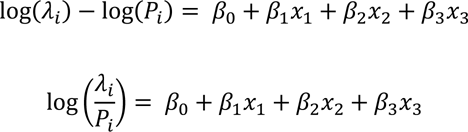

The multilevel model is formed by extending the underlying model to include two sets of random intercept terms, allowing for extra-Poisson variation at the MSOA level and for differences between the towns/cities/London Boroughs:

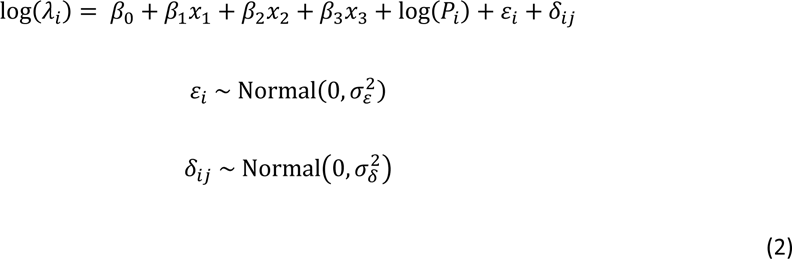

The two parameters, *ε*_*i*_ and *δ*_*ij*_, measure the deviation from the expected log infection rate at the MSOA and place levels. They measure the intra-place (within) and inter-place (between) differences, respectively. It is these that are used to form the index of exposure, measuring how exposed the average member of each ethnic group is to neighbourhoods with higher or lower than the expected infection rate.

The index is based on that used in studies of segregation where,

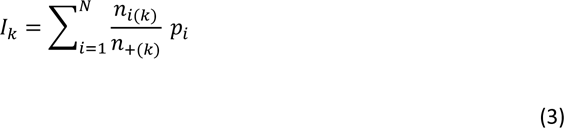

For such studies, *I*_*k*_ is the index value for ethnic group, *k*, of whom there are *n*_*i*(*k*)_ in neighbourhood, *i*, and *n*_+(*k*)_ for all *N* neighbourhoods within the study. The remaining value, *p*_*i*_, may be interpreted probabilistically. For a segregation index, it is the probability of selecting a member of a second ethnic group from the same neighbourhood that the member of ethnic group *k* is living in.^5^

The modification we make is that *p*_*i*_ becomes the probability of selecting, from a Normal distribution, a value lower than the MSOA’s deviance from its expected infection rate for the week. This means that if most of an ethnic group is living in MSOAs where the infection rate is higher than expected then the index will tend towards one, getting closer to one the greater the deviance from expectation. If most are living in neighbourhoods where the infection rate is lower than expected, it will tend towards zero. If the group is spread amongst neighbourhoods where those with higher infection rates balance out those with lower, then the index value will be 0.5.

Specifically, *p*_*i*_ is extracted from *ε*_*i*_ and *δ*_*ij*_ in Equation 2. These are treated as quantiles from a Standard Normal distribution and used to generate the probabilities that feed into the index of exposure. With reference to Equation 3, *p*_*i*_ = *P*(*z* < *ε*_*i*_ + *δ*_*ij*_) or *p*_*i*_ = *P*(*z* < *ε*_*i*_) where *z* represents quantiles from a Standard Normal distribution. Critically, the two variants of *p*_*i*_ mean that the index can be calculated with or without place effects, the latter controlling for the differences between towns and cities and therefore the broader scale geography of the disease. This is discussed further in the results section.

The model is fitted 49 times, once for each week of the pandemic included in the study. It follows that the index of exposure is calculated weekly too. Adding-in the subscript, *t*, to denote time gives,

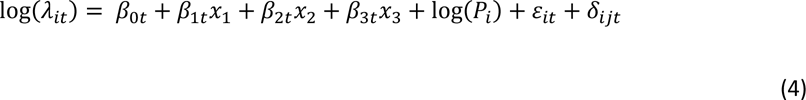

This shows that the modelled rate varies weekly as do the random intercepts but not the control variables because the population size, demographic profile and number of care homes beds are assumed to be constant for the study period. The regression coefficients could also be assumed to be time invariant, calculated once for the whole period and fixed at those values. However, such an assumption would be misplaced so their coefficients are permitted to vary too.

Although estimating their effect size is not the purpose of the model, their changing values are of passing interest and are shown in Figure 1. The number of care home beds per adult population had the greatest impact on increasing neighbourhood infection rates as the first national lockdown ended, in May 2020 (the periods of national lockdown are indicated in the figure with light grey shading). This is when concern about the lack of protection afforded to care home residents and their staff was of considerable media interest. Greater percentages of those aged 18-21 generally has a negative effect on the infection rates but not in the first week of October, in particular, when it had the strongest effect on increasing infection rates. This is when students returned from their homes to their term-time addresses after a delay in the start of the University term. The effect of populations aged 22-35 on infections is greatest after the first lockdown had ended and through the summer when pubs and restaurants had partially reopened and more people had returned to their workplaces.

**Figure 1.**
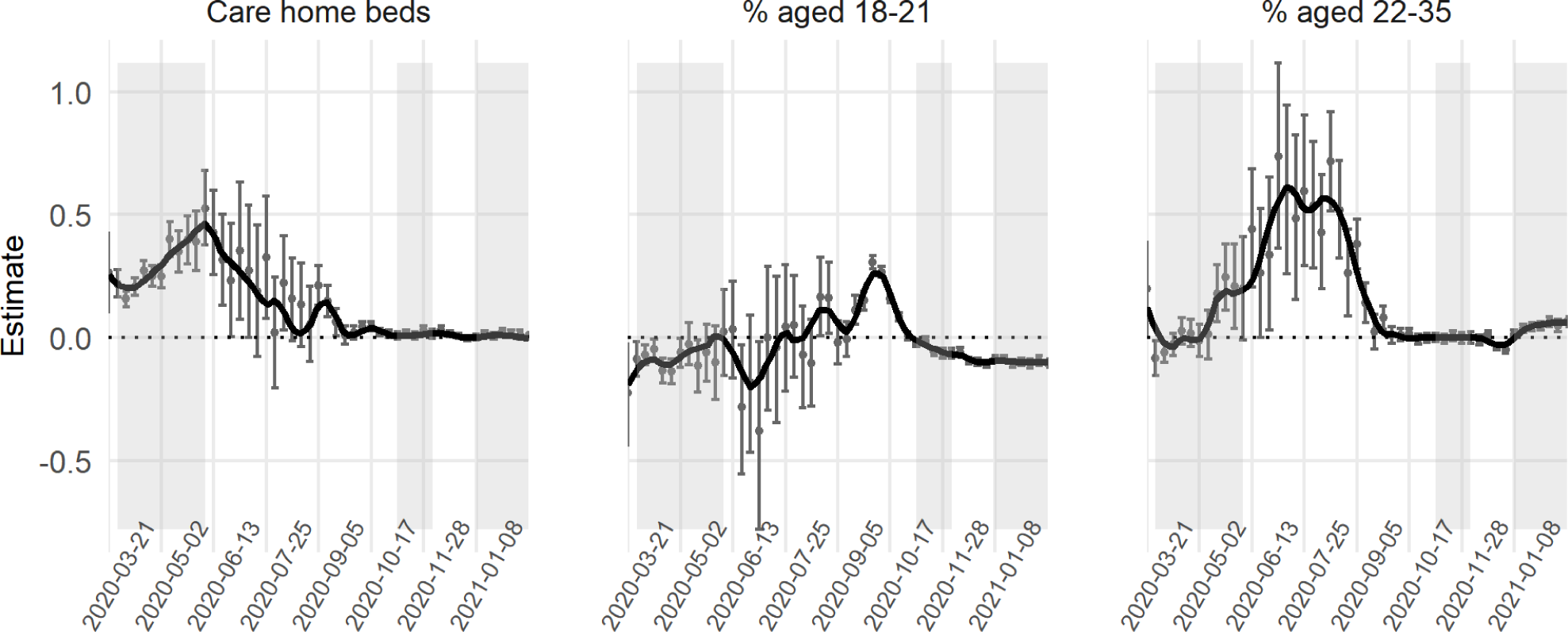
The regression coefficients for the control variables for each week of the study. The English national lockdown periods are shaded in grey.

The models are fitted as a Bayesian model using the brms package for R (Bürkner, 2017, 2018), which provides an interface to Stan (Stan Development Team, 2019). Results takes the form of draws from a Bayesian posterior distribution for the parameters of interest that are then summarised by their mean value.

## 4. Results

The index values, initially calculated with the inclusion of both the MOSA and place level effects, *ε*_*i*_ + *δ*_*ij*_, are shown in the upper part of Figure 2. Because the model is fitted separately for each week, the values are relative: they show which of the ethnic groups are living more or less in Covid-infected neighbourhoods that week, given the underlying infection rate.

**Figure 2.**
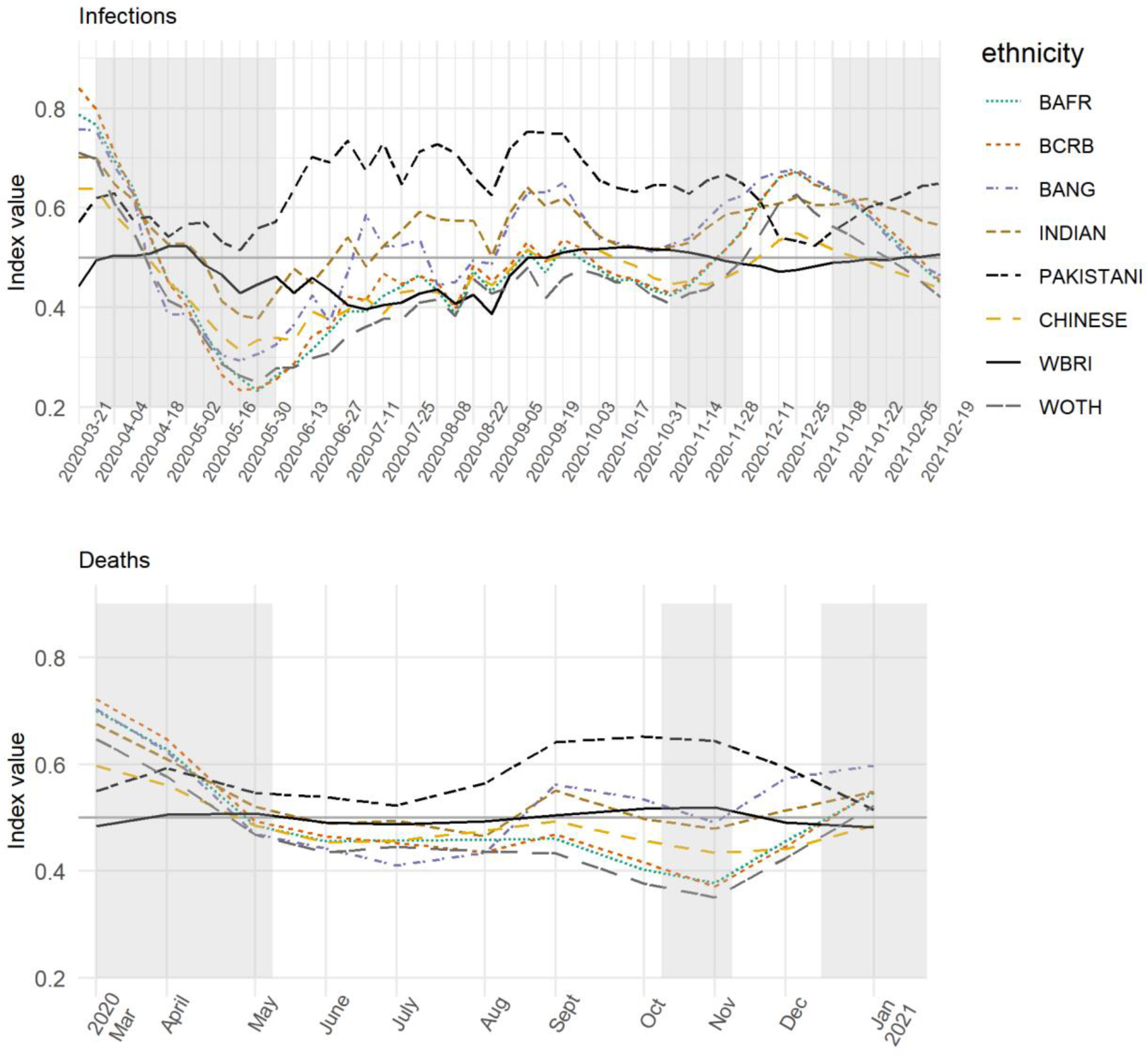
Showing (upper) the Index of Exposure of the various ethnic group to Covid-infected MSOAs for each week of the study, and (lower) recalculated for mortality rates. Note: BAFR = Black African; BCRB = Black Caribbean; BANG = Bangladeshi; WBRI = White British; WOTH = White Others.

To improve the legibility of the chart, some groups are omitted; specifically, the Arab, Black Other, Irish, Roma/Travellers, Other Asian, Other and joint ethnicity groups. The Black African group formed 1.8 per cent of the residential population of England in the 2011 Census; the Black Caribbean group, 1.1 per cent; the Bangladeshi, 0.8 per cent; Indian, 2.6 per cent; Pakistani, 2.1 per cent; Chinese, 0.7 per cent; White British, 85.4 per cent; and White Other, 4.6 per cent. Amongst those omitted, the Irish (1.0%), Other Asian (1.5%), Other (1.0%) and joint ethnicity (2.3%) groups formed a greater percentage of the Census population than some of those included but are removed either because they are less ‘distinct’ as a group or because they are not typically associated with BAME groups.

Figure 2 reveals that it was especially the Black groups that were most exposed to higher Covid-infected residential neighbourhoods earlier in the pandemic. However, this declines over much of the period of the study, rising again between the second and third lockdowns with the spread of the mutated virus in London and the South East. The implication is that when Covid-19 is spreading across the capital, it is the Black population that is most impacted. However, the group that is most consistently ‘over-exposed’ is the Pakistanis, with the highest index value for 38 of the 49 weeks (78%). The Indian and Bangladeshi groups often have higher index values too but usually not to the same extent.

Similar trends are found if the index is recalculated but modelling the numbers of Covid-19 deaths instead of infections. The results are in the lower part of Figure 2. The two sets of indices are not fully comparable, partly because the mortality data are monthly instead of weekly but more especially because they mortality index is modelled with the same care homes variable as for infections but also with the percentages of the adult population aged 25 to 29, 30 to 34, 35 to 39, and so forth to those aged 80 years or greater. The reason for including the additional age variables for mortality but not infections is that age does not cause infection but it is the greatest risk factor for death having been infected, especially for older populations. The additional co-variates act to age standardise the mortality data. Despite the model differences, it is again the Pakistani group with most frequently the highest index value, for 8 of the 11 months (73%).

We acknowledge that the ethnicity data underpinning Figure 2 are almost a decade old and that the population ‘as was’, at the time of the 2011 Census, will neither have remained in situ in each MSOA nor have been replaced on an exact like-with-like basis as people change their place of residence. It is known, from the segregation literature, that there has been a process of desegregation in most UK towns and cities as ‘minority’ groups have moved out from their traditional enclaves into what are becoming more mixed neighbourhoods (Johnston et al., 2013; Catney, 2015; Catney, Wright & Ellis, 2020). If that process has continued, which is likely (Harris & Johnston, 2020), and if, therefore, the Census data now overstate the geographical clustering of particular ethnic groups into particular neighbourhoods then the indices of exposure to Covid-19 could exaggerate the differences between the groups.

Unfortunately, there are few alternative sources of geographically detailed ethnicity data. Although consumer data hold promise (Lan, Kandt & Longley, 2020) they raise questions about their representativeness of the population and the biases they contain. In any case, it is highly unlikely that the members of the various ethnic groups have become sufficiently dispersed over the last nine years for the trends shown in Figure 2 to be only an artifact of dated information. For the groups to become that spread out would require the disappearance of the socio-spatial processes and spatial inequalities that led some groups to be more concentrated in some places than others – a farfetched proposition that is refuted by the evidence (Byrne, 2020).

The greater problem in interpreting Figure 2 is not the age of the ethnicity data but the changing regional geography of the disease over the study period. The higher exposure of Black groups in the initial period of the first wave is a function of what was, at that time, the higher prevalence of Covid-19 in London, where a greater share of the Black groups lives. The subsequent decline in their exposure, superseded by Asian groups, reflects the subsequent outbreaks in parts of the Midlands and more northerly regions. The later period, when the Pakistani group was briefly not highest again and less than the Black groups – towards the end of November 2020 through to January 2021 – reflects the emergence of the mutated strain of Covid-19 in London and the South East, prior to another national lockdown.

This changing geography of infections is revealed by plotting *δ*_*ij*_, the deviance, at the town and city level from the expected log infection rate for that week. The plots are in Figure 3, to which some smoothing has been applied to improve the visual clarity of the charts, drawing out the broad trends.

**Figure 3.**
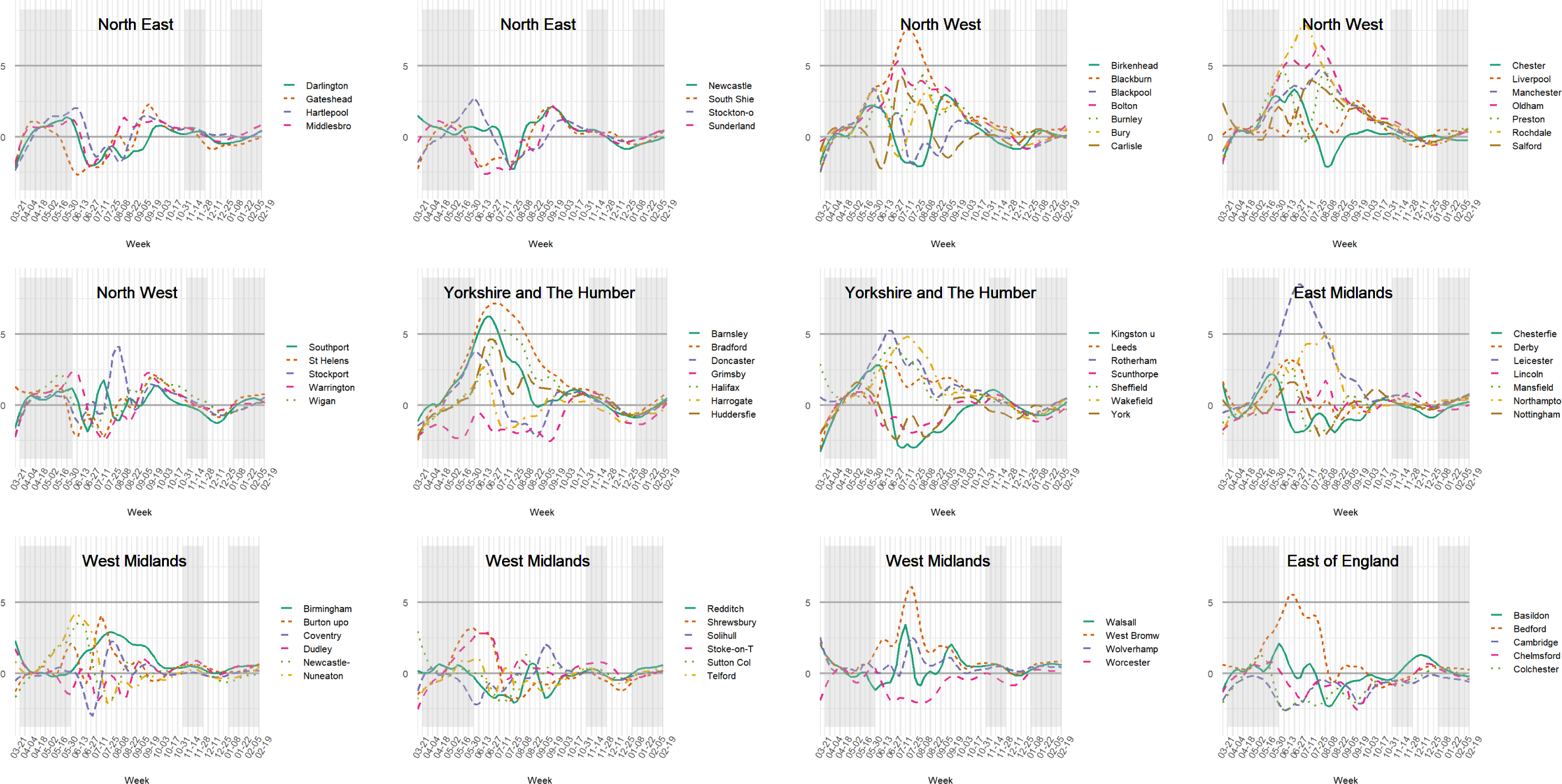
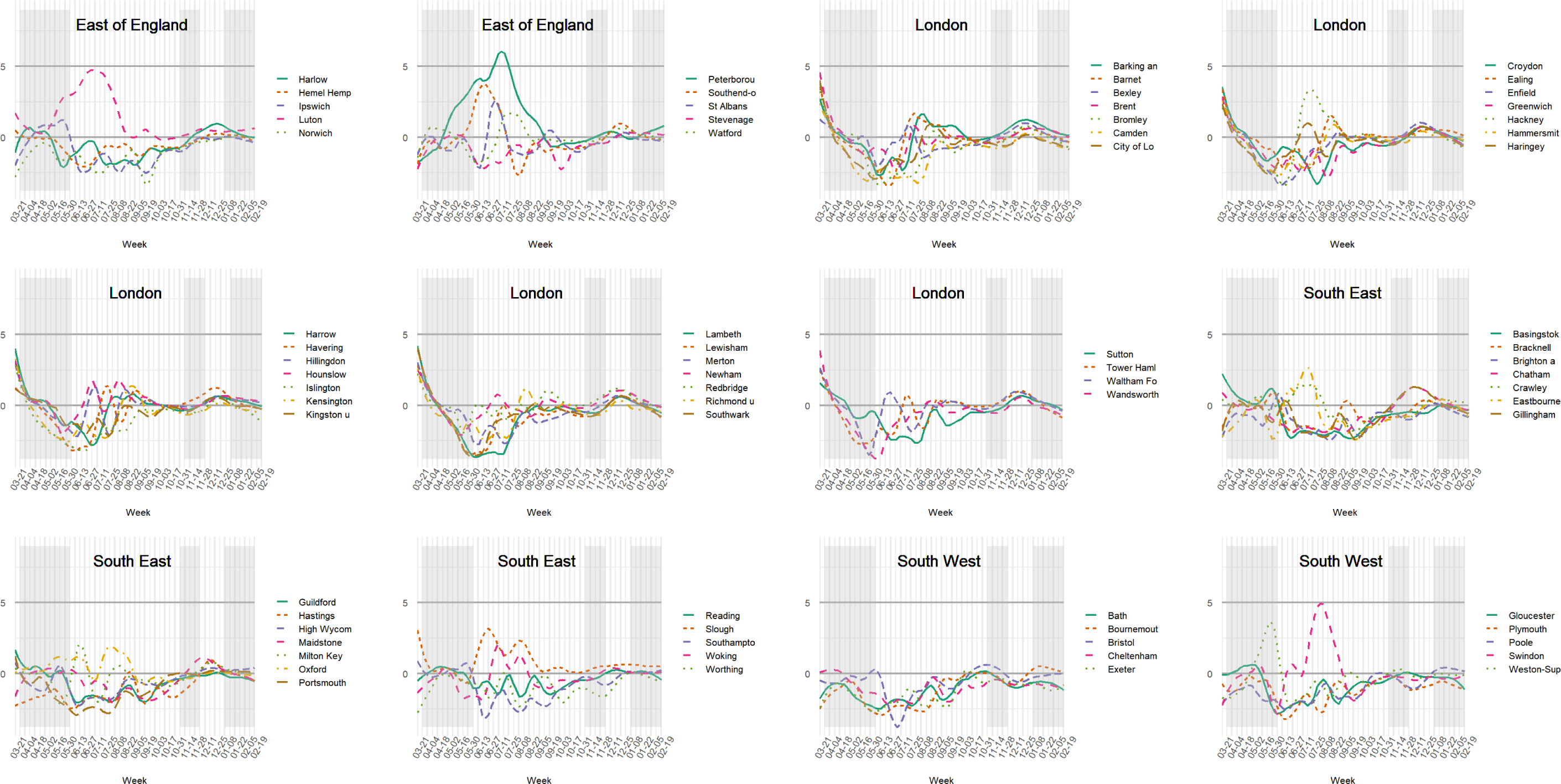
Indicating how much individual towns and cities in the English regions deviate from their expected log infection rate each week of the study. Smoothing has been applied and place names have been truncated in some cases.

London had higher than expected rates early-on but those quickly tailed off. In the middle period between the first two national lockdowns, the infection rates were higher than expected in parts of the North West, Yorkshire and The Humber, and in the East Midlands – in places such as Blackburn (about 27 per cent Indian, Pakistani or Bangladeshi in its 2011 Census population), Rochdale (13%), Liverpool (2%), Oldham (18%), Bradford (25%) and Leicester (32%). Most of these have comparable or greater percentages of the three Asian groups than London (12%), with the exception of Liverpool. By the end of the study period most places have converged around the zero line in Figure 2. This is not because the number of infections is reducing. Quite the opposite: it is because the disease has spread so widely that there is less variation between places.

These geographical variations are not unimportant because they invite the question of how they arose. It has been suggested that Londoners initially had higher relative exposure because theirs is a world city with greater travel connections both to the rest of the UK and across the world; also within London itself, with a much more extensive public transport system than other English towns and cities (Harris, 2020). But why did Leicester, for instance, have a period of greater infections necessitating a local lockdown? The answer is likely to lie within its borders – the housing stock, for example, or employment types, amongst other attributes of the city and its population. The place represents the spatial context and to discount that contextual variation could downplay the likelihood that what occurred in Leicester was not purely happenstance but reflects something about that city, to which Asian groups, amongst others, were exposed, increasing the infections.

Nevertheless, even within places where the overall infection rate is high, it is possible that some groups are living in neighbourhoods where the rate is even higher. Or, in places where the overall rate is low, still some groups face much higher exposure. It is therefore instructive to modify the index so that the broader scale differences between the towns and cities are removed, leaving the exposure to be measured relative to the local context. If it emerges that one or more groups are still more exposed to Covid-19 than others then it cannot be attributed only to the broad, regional geography of the disease. It means that the differences between the groups also emerge within individual towns and cities, not just between them.

The way to make the modification is to recalculate the index of exposure without the place effects from the underlying model. In terms of Equations 2 and 3, above, this means using only *ε*_*i*_, and not *ε*_*i*_ + *δ*_*ij*_, giving an index of local exposure. The results are shown in the upper part of Figure 4, drawn to the same scale as Figure 2 on the y-axis. The differences between the ethnic groups have reduced; unsurprisingly as we have removed the geographical variation between the towns and cities (but not within them). Whilst the remaining differences are not always large, it is the persistence of the difference between the Pakistani and other groups that remains of primary interest. In 47 of the 49 weeks (96%), the Pakistani group had the greatest local exposure, of which 43 weeks are consecutive.^6^ We believe that this finding should not be understated because even under a restricted scenario that focuses only on urban areas and which removes the broader-scale patterns in the disease *still* the Pakistani group is found to be more exposed to the higher Covid-infected neighbourhoods than are other groups. The Chinese and the White British who are often the least exposed groups, at the local level.

**Figure 4.**
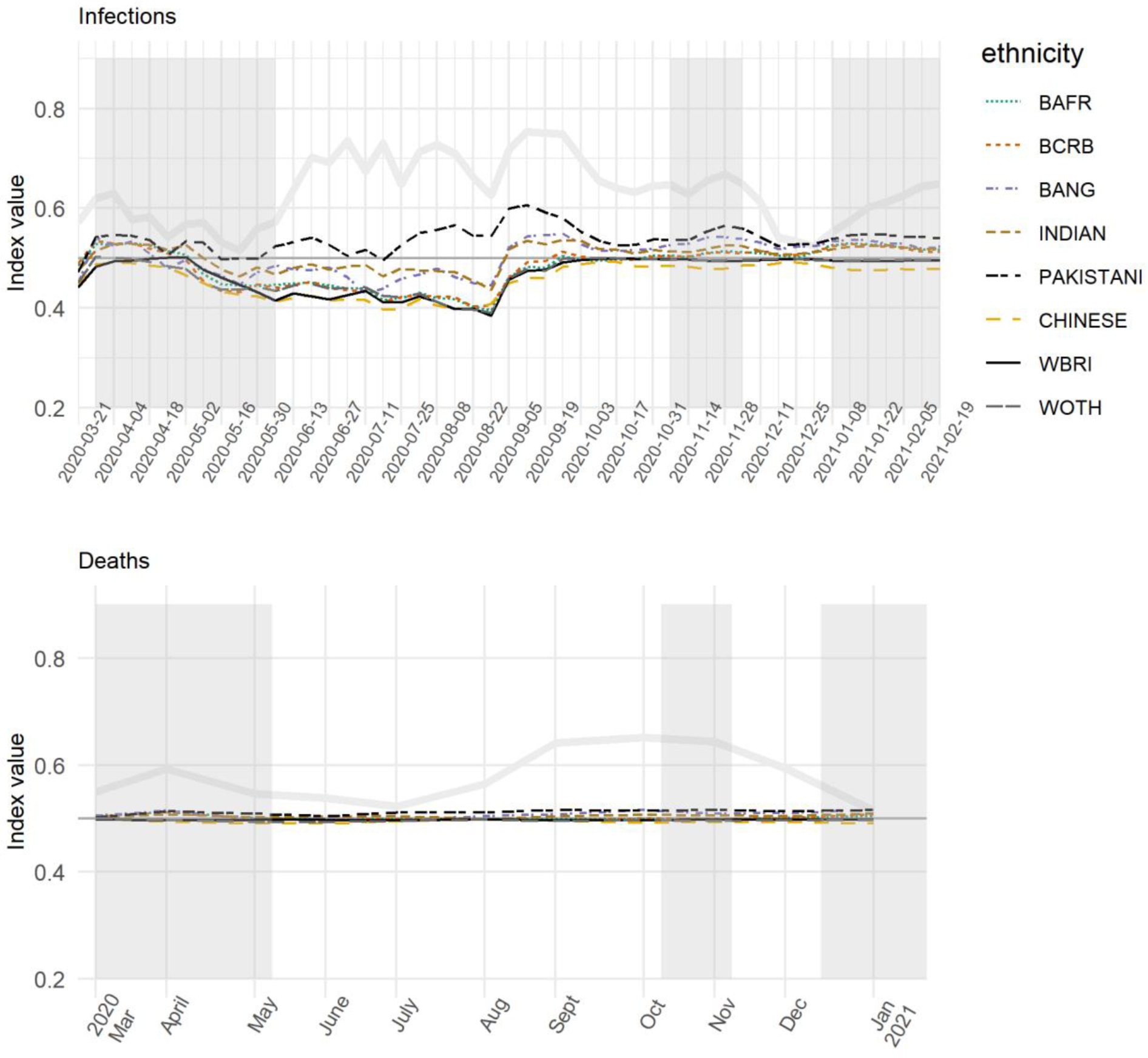
Showing the Index of Local Exposure values, which controls for the broader scale geography of the disease each week and is based on the variations within towns and cities rather than between them. The previous values for the Pakistani group are indicated in light grey.

However, the differences all but disappear between the ethnic groups when the index is recalculated for the age-standardised mortality rates, In fact, the Pakistani group does still have the highest value for 8 of the 11 months but the differences are trivial and illegible on the chart. The implication of this is that urban inequalities in mortality are more strongly expressed at the regional level and between major towns and cities (Griffith et al., 2021), whereas inequalities in infection are evident within towns and cities.

## 5. Discussion

The preceding analysis shows that some BAME groups have faced greater exposure to Covid-19 but not uniformly so and not always at the same time. Amongst them, it is the Pakistani and, to a lesser extent, the Bangladeshi and Indian groups that have had the greatest exposure to Covid-19 infected neighbourhoods within English major towns and cities. Their increased exposure partly relates to the overlapping geographies of the pandemic and of the residential geographies of where BAME live in the country. However, it is striking that for nearly every week of the pandemic the Pakistani group emerges as the one with the highest amount of local, neighbourhood exposure even after controlling for the variations between the towns and cities and therefore the broader scale geography of the disease.

Furthermore, the finding is not confined to what might be regarded as the most economically disadvantaged neighbourhoods because it is broadly consistent for areas of lowest and of higher average house price. To evidence this, Figure 5 shows the results from recalculating the index of local exposure with each of the ethnic groups split into ten sub-groups according to the decile of the trimmed mean house price of the properties sold in their MSOA between January 1, 2017 and February 27, 2020. The deciles are calculated on a town and city basis, in this case treating London as a whole. This means that those living in the highest decile are in MSOAs with the most expensive property, on average, for their town and city, not necessarily nationwide. This is to prevent the top decile being dominated by London.

**Figure 5.**
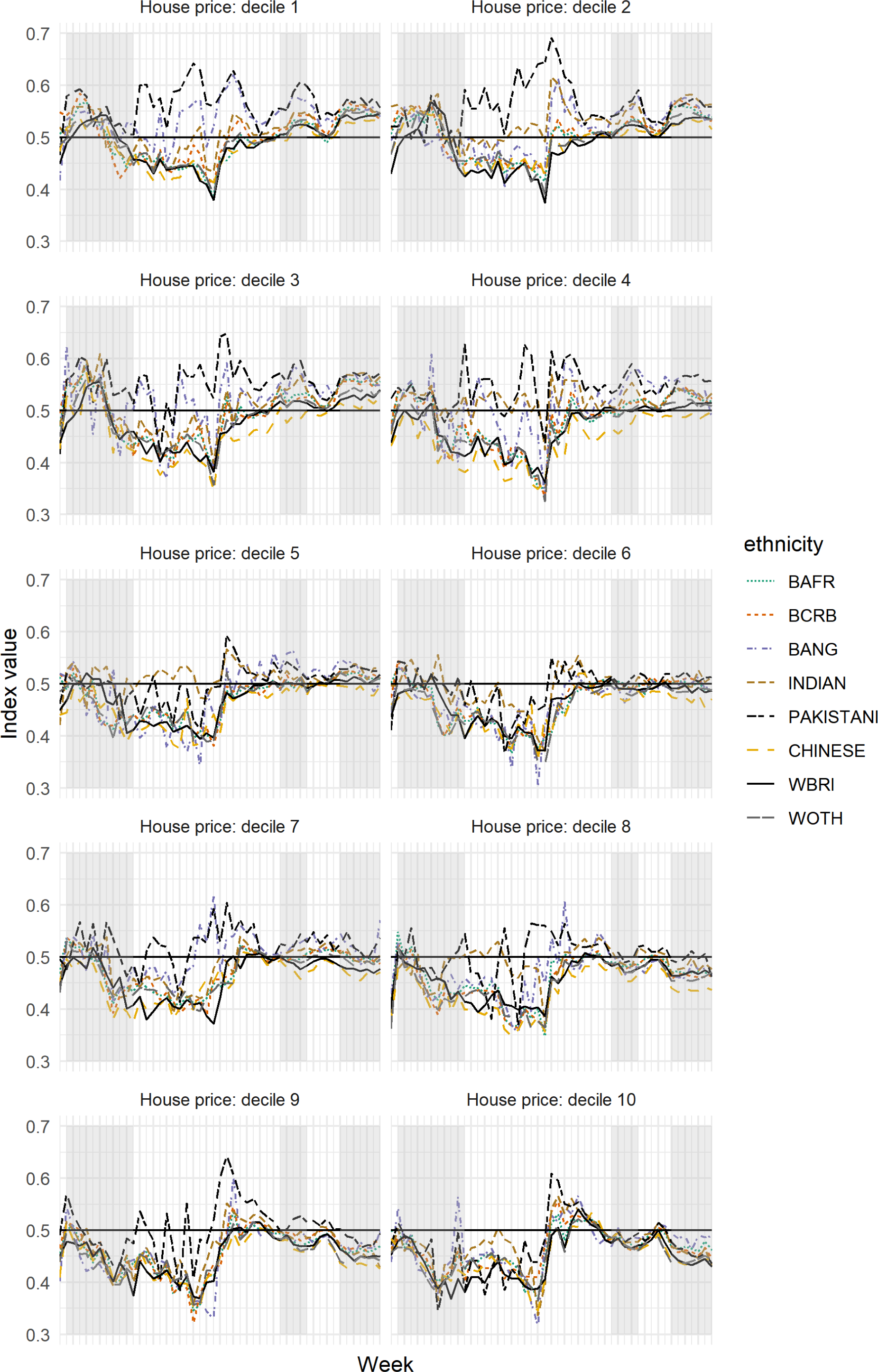
Showing the Index of Local Exposure values for each ethnic group, stratified by the average house price value of the areas in which they live. Decile 1 is the lowest average, decile 10 the highest, calculated relative to the town or city in which the properties were sold.

As before, it is not always the magnitude of the differences that are of most interest but the consistency of them disadvantaging the Pakistani group. This appears not to be a straightforward function of neighbourhood deprivation because the greater exposure of the Pakistani group to Covid-infected neighbourhoods is not restricted to the neighbourhoods of least cost housing. It is present in those neighbourhoods: for decile 1, the Pakistani group has the highest index value of all groups for 39 of the 49 weeks (80%). However, that number is exceeded by that for decile 9 – amongst the most expensive housing – for which the index value is highest for the Pakistani group for 40 of the weeks. Similar values are found for deciles 2 (29 weeks), 3 (37 weeks), 4 (32 weeks) and 7 (30 weeks). The numbers are lower for deciles 6 and 8 (23 and 28 weeks, respectively) but it is only for deciles 5 and 10 that Pakistanis are not the most frequently exposed group. For these two deciles, the frequency is exceeded by the Bangladeshi and Indian groups.

Why is the Pakistani group, especially, living in neighbourhoods with higher infection rates locally? *The Doreen Lawrence Review* identifies two key factors: environmental exposure – poor quality and overcrowded housing – and occupational exposure. Evidence for the first of these is suggested by looking at the residential density of each MSOA, calculated as the mid-2019 population estimate divided by the number of domestic mail delivery points per MSOA.^7^ For the average member of each ethnic group, the residential density is, in ascending order: 2.28 people per dwelling for the White British; 2.48 for the White Other group; 2.49 for the Chinese; 2.58 for Black Africans; 2.60 for Black Caribbeans; 2.67 for Indians; 2.72 for Bangladeshis; and, 2.79 for Pakistanis.

On average, therefore, members of the Pakistani group are living in higher residential density areas. Moreover, this result holds for nearly every one of the property price deciles: the residential density for the average Pakistani group member is greater than for any other group, except in deciles 5, 9 and 10. The densities are shown in Table 1, which also reveals that the residential density is always lowest for the average member of the White British group.

**Table 1.** The estimated residential density (people per dwelling) for the MSOA of the average member of each ethnic group, by the decile of the average house price of the MSOA.

| Decile | BCRB | BAFR | BCRB | BANG | INDIAN | PAKISTANI | CHINESE | WBRI | WOTH |
| --- | --- | --- | --- | --- | --- | --- | --- | --- | --- |
| 1 | 2.65 | 2.66 | 2.65 | 2.84 | 2.70 | 2.89* | 2.50 | 2.34# | 2.50 |
| 2 | 2.70 | 2.67 | 2.70 | 2.93 | 2.86 | 3.03* | 2.54 | 2.37# | 2.58 |
| 3 | 2.68 | 2.64 | 2.68 | 2.93 | 2.92 | 2.94* | 2.50 | 2.35# | 2.57 |
| 4 | 2.54 | 2.53 | 2.54 | 2.81 | 2.80 | 2.91* | 2.54 | 2.33# | 2.49 |
| 5 | 2.66 | 2.61 | 2.66 | 2.67 | 2.68* | 2.66 | 2.51 | 2.33# | 2.58 |
| 6 | 2.60 | 2.52 | 2.60 | 2.65 | 2.68 | 2.70* | 2.50 | 2.36# | 2.58 |
| 7 | 2.62 | 2.58 | 2.62 | 2.71 | 2.70 | 2.77* | 2.55 | 2.36# | 2.61 |
| 8 | 2.51 | 2.51 | 2.51 | 2.51 | 2.56* | 2.55 | 2.45 | 2.34# | 2.51 |
| 9 | 2.63 | 2.58 | 2.63 | 2.59 | 2.62 | 2.65* | 2.63 | 2.40# | 2.64 |
| 10 | 2.50 | 2.49 | 2.50 | 2.48 | 2.59 | 2.52 | 2.64* | 2.38# | 2.57 |

\* Highest value for the decile # Lowest value for the decile
\* Highest value for the decile

According to the English Housing Survey, April 2016 to March 2019, 18 per cent of Pakistani households were overcrowded, defined as a property having fewer bedrooms than it needs to avoid undesirable sharing, based on the age, sex and relationship of household members. That percentage is exceeded only for Bangladeshis, at 24 per cent. For the White British, it is 2 per cent (source: https://www.ethnicity-facts-figures.service.gov.uk/housing/housing-conditions/overcrowded-households/latest#by-ethnicity). ONS data from the Annual Population Survey household dataset, January to December 2018, show that 35 per cent of Pakistanis households are intergenerational households, with someone present from each of the age groups 0–19 years, 20 – 69 years, and aged 70 or over. Again, this is only exceeded by Bangladeshi households (56%) and lowest for the White British (1.5%) (https://www.ons.gov.uk/peoplepopulationandcommunity/birthsdeathsandmarriages/families/adhocs/12005householdsbyagecompositionandethnicityuk2018).

On face, there is not a strong relationship between residential density and how far each neighbourhood deviates, locally, from the expected, log infection rate. A simple linear regression suggests a Pearson correlation of *r* = 0.08, with *y*_*i*_ = *ε*_*i*_ as the dependent variable, the density values, mean-centred for each individual town and city, as the predictor variables, and with the separate weeks handled as fixed effects. This is a low effect size under Cohen’s (1988) criteria.

However, if the model is weighted according to the share of the total Pakistani population that lives in each neighbourhood then the correlation rises to 0.26, which is nearing a medium effect. This effect is greater for the Pakistani group then for any others. The corresponding correlation when weighting by the distribution of the Bangladeshi population is 0.17; by Indians, 0.16; by Black Caribbeans, 0.10; by Black Africans, 0.09; by White British and White Other groups, 0.06; and, by Chinese, only 0.04.

Similar analysis can be undertaken looking at the percentage of each neighbourhood’s residents in key worker jobs. Relevant data have been collated by Oxford Consultants for Social Inclusion (OCSI) from a 2018 Business Register and Employment Survey and are part of a larger dataset available at https://ocsi.uk/2020/04/01/covid-19-vulnerability-index-and-data-download/. The correlations between the local variations in the log infection rates and jobs in education, health, and transport and storage, are trivial: no more than *r* = 0.03 in each case. However, weighting by ethnicity gives correlations of 0.20 when weighting by the Pakistani group, 0.09 with the Bangladeshi group, 0.10 with the Indian groups, and values of between 0.02 and 0.06 for all other groups. These results suggest that the effects on Covid-19 infections of both occupational and environmental exposure are greater for the Pakistani group than for any other.

Bringing this together, the Poisson model of infection rates (Equation 4) is refitted to recalculate the local index of infection, this time conditional on additional co-variates for the log of the trimmed mean house price, mean-centred per town or city, residential density, the percentages of the neighbourhood populations employed as key workers, and the percentages of the adult population in various age groups from 18 to 80 and above. As previously, the coefficients for these variables are largely incidental to the analysis, although it may be noted that it is the percentage aged 70 to 79, residential density and average house price that have the greatest effect on the infection rates, also being significant, most often, at a 95 per cent confidence interval. Residential density is positively correlated with infections; house price and those in their seventies negatively so. Of interest, is what remains in the index of local exposure values, despite the additional co-variates, calculated from the remaining variations in the log infection rates within major towns and cities. Figure 6 reveals that the difference between the Pakistani and other groups has been reduced. However, the Pakistani group still has the highest value for 35 of the 49 weeks (previously it was 47), with the Indian group being highest for 9, the Bangladeshi group for 7, and the Black Caribbeans for 2.

**Figure 6.**
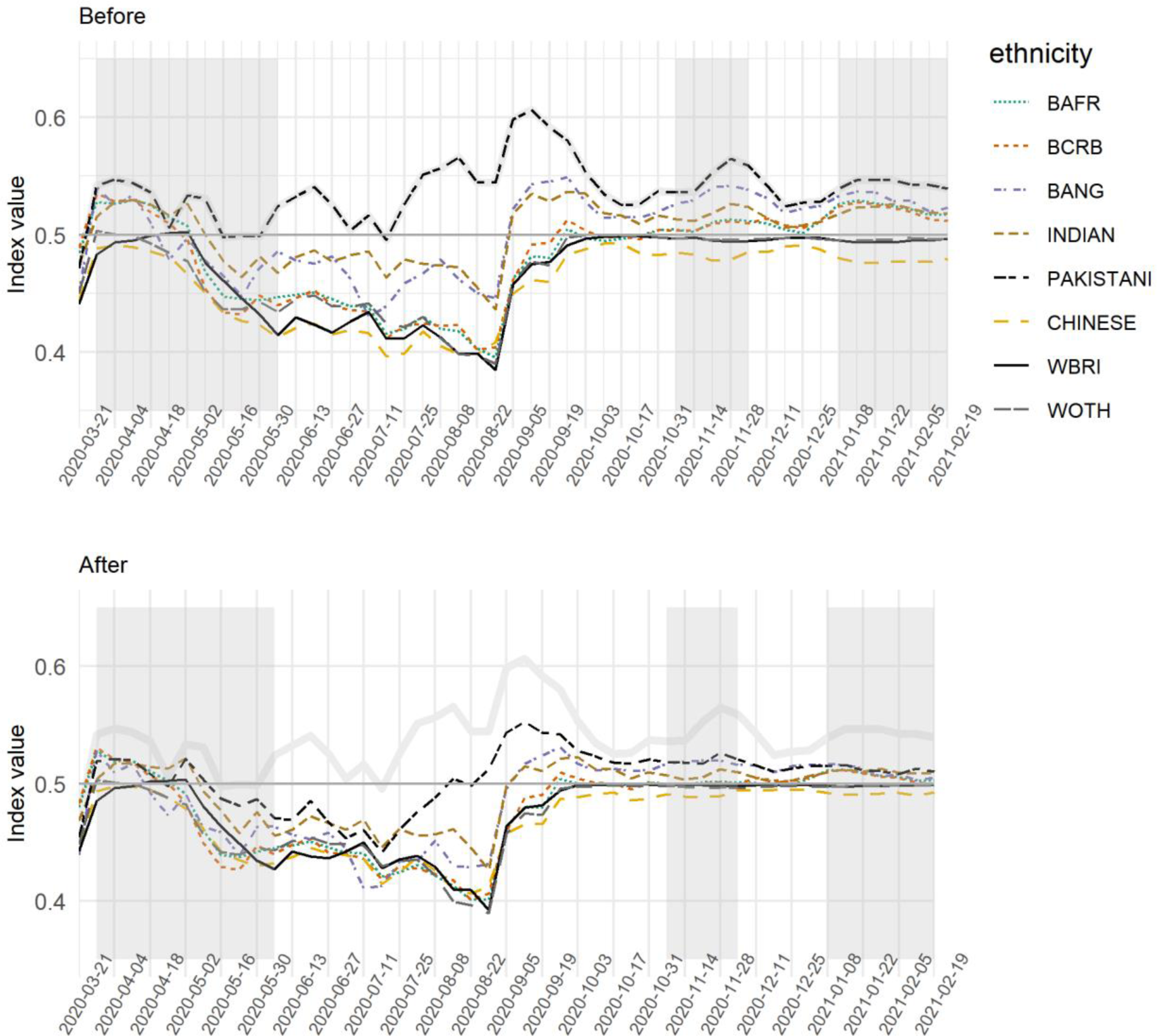
Showing the Index of Local Exposure values before and after the inclusion of the additional predictor variables.

## 6. Conclusion

This paper has developed an index of exposure using an underlying Bayesian Poisson model of the Covid-19 infection rates for neighbourhoods in English towns and cities. The index measures how exposed various ethnic groups, including BAME groups are to Covid-infected residential neighbourhoods and can be modified to allow for the broad-scale geography of the disease, focusing, instead, on the localised highs and lows and who is exposed to them.

The results show that members of BAME groups tend to be living in neighbourhoods where the infection rates are higher, reflecting their greater risk of being infected by and dying from the disease. The higher index values are true of Black groups in the initial weeks of the pandemic when the rates of infection are highest in London but become more greatly characteristic of Pakistani and, to a lesser extent, Bangladeshi and Indian groups as the prevalence of the disease shifts to other towns and cities that include Oldham, Bradford, Blackburn, Manchester, Rochdale, Northampton and Leicester.

On February 18, 2021, the Department of Health and Social Care published interim findings from a study, showing that “large household size, living in a deprived neighbourhood, and areas with higher numbers of Asian ethnicity individuals were associated with increased prevalence” (Department of Health and Social Care, 2021). This accords with our own findings, with the caveat that we observe the higher exposure values for the Pakistani group not solely to be associated with the least wealthy neighbourhoods but remaining evident when the data and the index are stratified into deciles based on property prices. This may be because the neighbourhoods concerned, the Middle Level Super Output Areas, are internally heterogenous regarding their housing stock, especially within major towns and cities. In any case, we agree with *The Doreen Lawrence Review* in citing environmental exposure as a concern. Ethnic inequalities in housing mean that members of the Pakistani group are living in higher residential density areas and, with the exception of the Bangladeshi group, are more likely to be living in overcrowded and/or intergenerational households.

Shankley and Finney (2020) observe that ethnic inequalities in housing “stem from the particular settlement experiences of postwar migrants to the UK in terms of the locations and housing areas afforded to them […] consolidated by dramatic changes to the UK’s housing landscape over recent decades [that have] exacerbated housing disadvantage for minorities” (p.149). They note the practices of discrimination and racism that existing in housing, the reduction in the social housing stock available and problems of affordability and financial risk – the overarching issue of excessive housing (including rental) costs, relative to income, in many English towns and cities that adds to financial pooling and sharing, and to overcrowding.

Housing inequalities are not the only explanation for why particular ethnic groups live, disproportionately, in Covid-infected neighbourhoods. The *Doreen Lawrence Review* also identifies occupational exposure, which we find evidence for too. Both are linked to other sub-national, socio-economic inequalities including the imbalanced nature of regional economies and employment structures in the UK (ONS, 2020d). Writing in *The Guardian*, Dorling (2020) picks up these themes, arguing that,

> in poorer, more often northern, parts more people have jobs that cannot be done from home and more use public transport. Frequently, childcare is provided by the extended family who live nearby – wages and benefits are usually too low to allow other childcare options. There is less early retirement and more pensioners need to work too. Further, overcrowding in homes in cities is more common and anyone out of work exacerbates that.

Although there is an element of caricature there, the broad impression helps inform Dorling’s conclusion that the key to understanding the geography of Covid-19, “is the underlying social and economic geography of England. To understand the changing medical geography of this pandemic, you must first understand how the country lives and works.” We agree.

## Data Availability

Please contact the authors

## Footnotes

1 A reason for the usually higher mortality amongst White groups is that the White British population is, on average, older.

2 Infection rates were also at their highest during the second wave, peaking, at a UK rate of 600 cases per 100,000 at the end of the first week of January.

3 The largest MSOA, by population size, is in Newham, London, with an estimated population of 26,513, of which 55 per cent are adults aged 22 to 35. This MSOA incorporates the Olympic Park and Stadium, including the Olympic Village, now converted to residential housing.

4 https://geoportal.statistics.gov.uk/datasets/major-towns-and-cities-december-2015-names-and-codes-in-england-and-wales

5 It is *p*_*i*_ = *n*_*i*(*l*)_⁄*n*_*i*+_ where *n*_*i*+_ is the total population of *i* and *l* denotes the second group: see, *inter alia*, the appendix of Harris & Johnston (2020), or Kaplan (2018).

6 If the figure is redrawn with some smoothing and a confidence interval for each group, then that for the Pakistani group typically overlaps with few or no other others each week suggesting it is improbable that the persistence of the Pakistanis as the most exposed group is of no statistical or substantive significance.

7 The delivery point information is from Ordnance Survey Code-Point data

